# A proteogenomic approach to identifying a gene signature associated with *HOXB13* G84E carrier status in prostate cancer tumours

**DOI:** 10.1101/2025.07.08.25331050

**Authors:** Kelsie Raspin, Zainab Noor, Adel T Aref, James R Marthick, Chol-hee Jung, Shaun Donovan, Steve Williams, Peter G Hains, Phillip J Robinson, Rosemary L Balleine, Qing Zhong, Roger R Reddel, Liesel M FitzGerald, Joanne L Dickinson

## Abstract

The rare *HOXB13* G84E variant is one of few established high-risk prostate cancer variants, yet the underlying molecular mechanisms by which it drives tumour development remain poorly understood. Prostate tumours from six G84E variant carriers and 18 non-carriers (wildtype) were examined using the Human AmpliSeq Transcriptome Gene Expression Panel and mass spectrometry. We identified a 799-gene expression profile and a 75-protein expression profile in G84E positive tumours when compared to wildtype tumours. Cluster analysis revealed that the top cluster consisted of 23 proteins which mapped to several lipogenesis pathways. Integration of the transcriptome and proteome data revealed overlapping differential expression of three genes (*NCLN, NFIB* and *VASN*), two of which have an established link to lipid metabolic pathways. Examination of publicly available transcriptome data from 493 TCGA primary PrCa tumours revealed 52 tumours with a similar pattern of differential expression of these three genes. We provide the first evidence from patient samples that the G84E variant is associated with upregulation of genes in the steroid metabolic, lipid biosynthesis and fatty acid metabolic pathways. We are also the first, to our knowledge, to use a proteogenomic approach to identify key molecular changes in prostate tumours from any high-risk rare variant carriers.

## 1. BACKGROUND

Prostate cancer (PrCa) has the highest heritability of any common cancer (1). Except for rare variants in DNA damage repair genes, including *BRCA1* and *BRCA2*, the rare G84E variant in *HOXB13* (rs138213197) remains the single most strongly associated PrCa risk variant. Originally identified in a familial PrCa cohort (2), this variant is associated with a 3-16-fold increase in PrCa risk across multiple studies, including our Tasmanian resource (OR=6.59; p=4.2×10^−5^) (3). The G84E variant is associated with early-onset familial disease (4–9), however there is no established association between carrier status and clinicopathological characteristics, including Gleason score (2, 6, 10, 11).

*HOXB13* is a transcription factor that is important for normal prostate development(12, 13), and is the only *homeobox* gene that maintains expression into adulthood (2, 12, 14, 15). It has been postulated that *HOXB13* can act as both an oncogene and tumour suppressor during carcinogenesis, and its role seems to be dependent on the cell environment. *HOXB13* plays a crucial role in the regulation of androgen receptor (AR) activity, a major regulator of the normal prostate and PrCa (16). The G84E variant is located within the MEIS binding domain of *HOXB13*, which interacts with the histone deacetylase, HDAC3. Recently, Lu *et al*., 2022 identified HDAC3 as a key cofactor of *HOXB13* and showed that together they reprogramme the epigenome and suppress the lipogenesis pathway (17). However, cell culture studies have indicated that the interaction between *HOXB13* and *MEIS1* is not impacted by the G84E variant (18). The exact role of the variant remains unclear, and it is still not known whether prostate tumours harbouring a G84E variant represent a distinct subgroup of tumours.

There is growing evidence that germline genetic variants, including rare variants, drive transcriptome and proteome signatures in tumours (reviewed in Sadeesh *et al.,* 2021 (19)). Defining ‘omic’ signatures can stratify patients, reveal molecular drivers of disease, inform response to tailored therapies and aid in the development of new therapeutics (20, 21). An integrated multi-omic approach has recently identified an 18-protein signature that is prognostic for risk of biochemical recurrence in intermediate-risk PrCa patients (22). Thus, multi-omic analyses can increase our understanding of disease pathogenesis and have the potential to advance precision medicine in the clinical setting. Moreover, most successful cancer therapeutics target protein functions (23). Therefore, a greater understanding of how the G84E variant impacts proteomic features may yield insights that will inform development of novel therapies. In the study reported here, we aimed to determine whether prostate tumours from *HOXB13* G84E rare variant carriers have distinct proteome and transcriptome signatures. We hypothesised that a proteogenomic approach could identify key signatures and pathways associated with the G84E variant, potentially highlighting future areas of researching focussing on novel therapeutic targets in carriers.

## 2. MATERIALS AND METHODS

### 2.1 Study Resources

This study utilises two Tasmanian PrCa cohorts: the *Tasmanian Familial Prostate Cancer Cohort* and the population-based *Tasmanian Prostate Cancer Case-Control Study* (previously described in ^3,33,45^). In total, 26 formalin-fixed, paraffin-embedded (FFPE) prostate tumour samples from 24 PrCa cases were available for this study. Two prostate tumour samples were available for two individuals (PC72-04 and PC72-06). The 24 cases included 20 men from the *Familial Cohort* and four from the *Case-Control Study*, comprising six *HOXB13* G84E variant carriers (all familial PrCa) and 18 non-carriers. Sectioned FFPE pathology blocks were histologically reviewed by a pathologist to provide a contemporary grading of the tumours (SD) and mark regions of malignancy (SD and RB). Ethics approval was obtained from the Human Research Ethics Committee Tasmania, Australia (H0017040) and written informed consent was obtained from all living participating individuals. For deceased cases, a waiver of consent was obtained to collect prostate tumour samples from pathology laboratories.

### 2.2 Nucleic Acid Extraction and Transcriptome Assays

Twelve of the 26 tumour blocks had sufficient malignant RNA for transcriptome assays. RNA and DNA were coextracted from FFPE tissue block sections, with regions of malignant prostate cells macro-dissected and extracted separately using the AllPrep DNA/RNA FFPE Kit (Qiagen). RNA was quantitated using the Agilent TapeStation System and DNA using the Nanodrop ND-1000 UV-vis spectrophotometer (Nanodrop Technologies). *HOXB13* G84E carrier status was confirmed by Sanger sequencing tumour DNA.

#### 2.2.1 AmpliSeq^TM^ Transcriptome Assays

RNA was assayed on the AmpliSeq^TM^ Transcriptome Human Gene Expression Kit (ThermoFisher Scientific) and sequenced on the Ion Torrent Proton^TM^ sequencing system, as previously described ^44,46^. The Ion Torrent Suite v5.02 software was used to map the reads and reads per million (RPM) values were calculated.

#### 2.2.2 Data Pre-Processing for Differential Gene Expression Analysis

Gene expression values of the 20,810 genes captured by the AmpliSeq^TM^ Kit were log2 transformed and standardised to the z-score (Supplementary Figure 1). In short, z-score normalisation was performed across all genes so that relative expression levels of each gene could be assessed between all samples. There were 860 genes in the dataset that had either no expression values in all of the samples or had values in only a single sample, hence these genes were removed from the data matrix. Genes with missing data in >50% of the samples were removed, leaving 17,128 genes for differential gene expression analysis.

### 2.3 Protein Extraction and Mass Spectrometry

Microtome sections from 26 prostate tumour samples were prepared as previously described ^24^. All samples were processed at the same time to avoid batch effects and the data were acquired on two mass spectrometers. HEK293 digestion controls were included. An Eksigent nanoLC 425 HPLC (Sciex) was coupled to a 6600 Triple TOF (Sciex) mass spectrometer. The data independent acquisition mass spectrometry (DIA-MS) details were as described by Lucas *et al.,* 2019 ^24^. See Supplementary Information for variable windows utilised. MS runs were randomised and run in technical duplicates. Each sample was spiked with unlabelled, non-human peptides as retention time (RT) standards.

#### 2.3.1 Spectral Library Generation and SWATH Data Extraction

MS data were searched using DIA-NN 1.8 ^47^. Canonical SWISS-PROT entries were extracted from Uniprot for *Homo sapiens*. The spectral reference library (SRL) was constructed using all SWATH runs and the *Homo sapiens* FASTA file with DIA-NN. The final SRL, used for searching the data, contained 4,547 proteins and 21,422 peptides.

For SWATH data extraction, DIA-NN was employed using RT-dependent normalisation and data were extracted for 26 samples from 24 PrCa cases. Samples were run in technical duplicates with a total of 50 SWATH runs (two samples had only one replicate). In addition to patient samples, six HEK293 control runs were also added to the dataset. Therefore, a total of 56 SWATH runs were searched using the SRL.

#### 2.3.2 Data Pre-Processing for Differential Protein Expression Analysis

From DIA-NN search results, the precursor data were filtered for proteotypic peptides only with Global.Q.Value < 0.01. Proteins were then quantified using maxLFQ, with default parameters ^48^ and implemented using the DIA-NN R Package (https://github.com/vdemichev/diann-rpackage). A total of 14,908 peptides corresponding to 3,180 proteins were quantified. The quantified proteins ranged from approximately 2,000 to 3,000 proteins per sample. When evaluated, a missing rate of ∼10% to ∼70% per sample was observed, with missingness variable across instruments (Supplementary Figure 2), however this effect was reduced after normalisation (Supplementary Figure 3). Proteins with missing data in >50% of the samples were filtered (n=751) and the technical reproducibility of samples was evaluated by the Pearson correlation coefficient (Pearson’s r) among sample replicates. A high degree of correlation was observed, ranging from 0.83 to 0.91. Prior to downstream data analysis, data were log2 transformed, and sample replicates were merged to generate the final protein matrix. Where two prostate tumour samples were available from the one individual (PC72-04 and PC72-06), the sample with the highest percentage of viable cancer was included in subsequent analyses. Out of 3,180 proteins, 1,250 proteins were quantified in >90% of the samples and 140 proteins were quantified in <20% of the samples (Supplementary Figure 4), leaving 2,429 proteins for analysis in 23 prostate tumour samples (Supplementary Figure 5).

### 2.4 Differential Expression Analysis

Gene and protein expression data were standardised to z-scores. To determine differences in expression between *HOXB13* G84E variant positive and wildtype tumours, the Empirical Bayes moderated t-statistic in the *limma* R package (version 3.54.1) was used to calculate p values. Limma assumes a t-distribution for the test statistic, and the empirical Bayes moderation makes the estimate more robust. Significant DEGs and DEPs were selected using a raw p-value <0.05 and a log fold change (expressed as the difference in the group means) cut-off of ±0.5. Volcano plots were generated using the R package, *ggplot2* (version 3.4.1) and heatmaps were generated using the R package, *pheatmap* (version 1.0.12). For heatmaps, the complete linkage clustering algorithm was used, along with Euclidean distance as the distance measure.

### 2.5 Functional Enrichment Analysis

GO enrichment analysis of the DEGs and DEPs was performed using the PANTHER19.0 Classification System ^49,50^ available through Gene Ontology Unifying Biology (https://geneontology.org/docs/go-enrichment-analysis/, release 2024-11-03) ^51,52^. Cluster analysis to predict protein-protein interactions was performed using STRING (https://string-db.org/, version 11.5). The type of interactions between proteins was indicated by the network connection line, as determined by curated databases or experimentally. The input parameters were p-value <0.05, minimum gene count of 3, and an interaction score of 0.4 (and an enrichment score >1 for the protein data). kmeans clustering was used to identify the top two clusters from each analysis.

### 2.6 The Cancer Genome Atlas dataset

TCGA dataset utilised in this study consisted of somatic data from 493 radical prostatectomy prostate adenocarcinoma samples. The data was accessed through the Pan-Cancer Atlas (PanCanAtlas) initiative ^53,54^, as previously described ^46^. Here, mRNA expression profiles (batch normalised from Illumina HiSeq_RNASeqV2) of *NCLN, NFIB* and *VASN* were sourced through the cBioPortal for Cancer Genomics (https://www.cbioportal.org/datasets). Here, the median expression value of each gene was determined, and tumours with reduced expression of *NCLN, NFIB* and *VASN* (in comparison to the median) were identified. These tumours were determined to have a G84E-like expression signature. Clinicopathological features of disease, including Gleason score and age at diagnosis, as well as *HOXB13* and *AR* gene expression and *HOXB13* methylation (HM27 and HM450 merge) were statistically assessed between the tumours with the G84E-like expression signature and the remainder of the dataset. P values <0.05 were considered statistically significant.

## 3. RESULTS

### 3.1 Gene expression analysis of *HOXB13* G84E variant positive and wildtype prostate tumours

Array-based transcriptome assays were performed for 12 prostate tumours from the Tasmanian studies, including five *HOXB13* G84E positive and seven wildtype tumours (Table 1; Supplementary Figure 1). We have previously shown that *HOXB13* expression levels do not differ between G84E variant and wildtype tumours (3) and results from this current study confirmed these findings (p=0.09; Figure 1A). The objective of this study was to identify molecular pathways that distinguish *HOXB13* G84E variant positive from *HOXB13* wildtype tumours.

**Figure 1.**
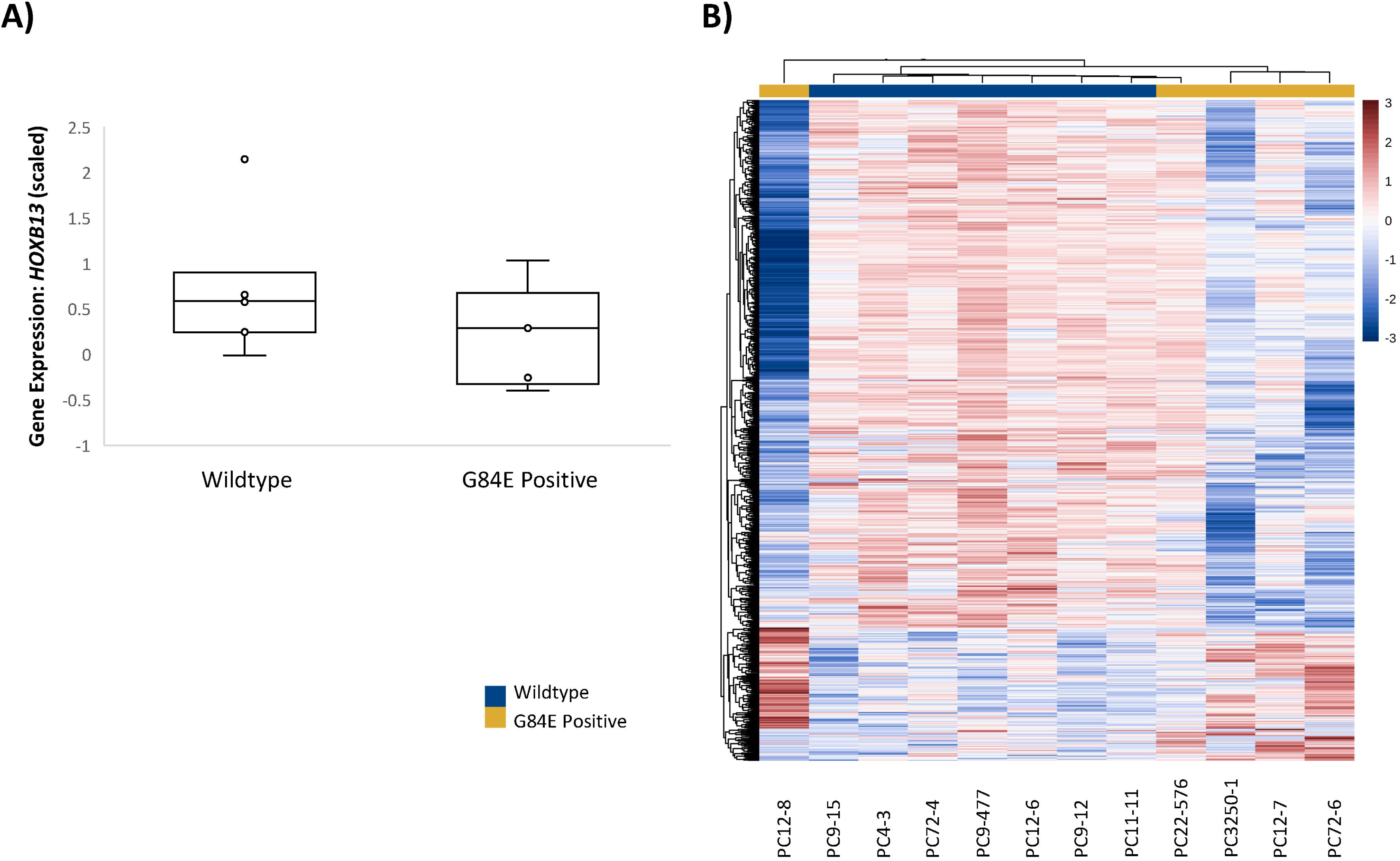
**A)** *HOXB13* gene expression (scaled) of G84E positive (n=5) and wildtype tumours (n=7, p=0.09). Individual samples are labelled. **B)** Heatmap of the 799 genes found to be differentially expressed between G84E positive and wildtype tumours.

**Table 1.** Clinical characteristics of prostate tumour samples available for this study.

| Sample ID | Age at Diagnosis (Range) | G84E Tumour Genotype | Tissue Source | Tumour Grade<sup>1</sup> | Gleason Score<sup>2</sup> | % of Viable Cancer | Malignant Tissue Available for Transcriptome Assays<sup>3</sup> |
| --- | --- | --- | --- | --- | --- | --- | --- |
| **G84E non-carriers** |  |  |  |  |  |  |  |
| DVA216 | 61-65 | CC | RP | - | 5 (3+2) | 20-50 |  |
| DVA220 | 61-65 | CC | RP | - | 6 (3+3) | <20 |  |
| DVA422 | 61-65 | CC | RP | - | 7 (3+4) | 20-50 |  |
| DVA1002 | 61-65 | CC | RP | - | 6 (3+3) | <20 |  |
| PC4-3* | 76-80 | CC | TURP | M/PD | 7 (4+3) | 50-80 | Y |
| PC9-7 | 71-75 | CC | TURP | - | 9 (5+4) | 50-80 |  |
| PC9-12 | 66-70 | CC | RP | - | 6 (3+3) | <20 | Y |
| PC9-15 | 61-65 | CC | TURP | - | 9 (4+5) | 80^^ | Y |
| PC9-158 | 61-65 | CC | RP | - | 6 (3+3) | <20 |  |
| PC9-477 | 51-55 | CC | RP | - | 6 (3+3) | 0^ | Y |
| PC9-627 | 61-65 | CC | RP | - | 7 (4+3) | 20-50 |  |
| PC11-11* | 81-85 | CC | TURP | - | 7 (3+4) | 20-50 | Y |
| PC12-1* | 61-65 | CC | RP | MD | 6 (3+3) | <20 |  |
| PC12-6* | 76-80 | CC | TURP | PD | 7 (3+4) | <20 | Y |
| PC12-9* | 66-70 | CC | TURP | - | 6 (3+3) | <20 |  |
| PC22-17 | 56-60 | CC | RP | - | 6 (3+3) | <20 |  |
| PC60-1* | 56-60 | CC | TURP | WD | 6 (3+3) | <20 |  |
| PC72-4* | 66-70 | CC | TURP | PD | 9 (4+5) | >80# | Y |
| **G84E variant carriers** |  |  |  |  |  |  |  |
| PC12-3* | 61-65 | CT | TURP | WD | 4 (2+2) | 0^ |  |
| PC12-7* | 56-60 | CT | TURP | PD | 9 (4+5) | 50-80 | Y |
| PC12-8* | 71-75 | CT | TURP | - | 6 (3+3) | <20 | Y |
| PC22-576* | 66-70 | CT | RP | M/PD | 7 (3+4) | <20 | Y |
| PC3250-1* | 51-55 | CT | RP | PD | 9 (4+5) | >80 | Y |
| PC72-6* | 61-65 | CT | TURP | W/MD | 10 (5+5) | 50-80<sup>#</sup> | Y |
| * Tumour included in our previously published work (3); <sup>1</sup>Tumour grade obtained from pathology report; <sup>2</sup>Contemporary Gleason Score from FFPE tissue block chosen for macro-dissection; TURP: Transurethral resection of the prostate; RP: Radical prostatectomy; WD: well differentiated; MD: moderately differentiated; PD: poorly differentiated; -: information was absent in original pathology report; ^Proteome removed from differential protein expression analysis due to no cancer present in tissue section; ^^Sample failed QC; <sup>#</sup>Two samples were assayed for this individual. Clinical data is representative of the prostate tumour sample with the highest % of malignancy, and the one included in subsequent differential expression analyses;<br><sup>3</sup>Malignant RNA was macro-dissected, extracted and assayed separately to benign prostate cells. |  |  |  |  |  |  |  |

Analysis of the 17 128 quantified genes that passed quality control (QC), revealed 799 genes that were differentially expressed between G84E variant carriers and non-carriers (Figure 1B), of which 159 were upregulated and 640 downregulated in G84E positive compared to wildtype tumours. A full list of the DEGs is provided in Supplementary Tables 1 and 2.

GO analysis of the 799 genes identified five distinct pathways, including protein localisation to organelle (GO:0033365), intracellular transport (GO:0046907), protein transport (GO:0015031), cellular biosynthetic process (GO:0044249) and protein metabolic process (GO:0019538; Supplementary Table 3). Cluster analysis found that the top cluster of genes (n=297) were involved in nucleic acid metabolic processes and transport, such as RNA and mRNA metabolic processes, translation and RNA splicing (Figure 2A; Supplementary Table 4). The second cluster included 229 genes (Figure 2B), 30 of which mapped to the cellular lipid metabolic process (Figure 2C; Supplementary Table 4).

**Figure 2.**
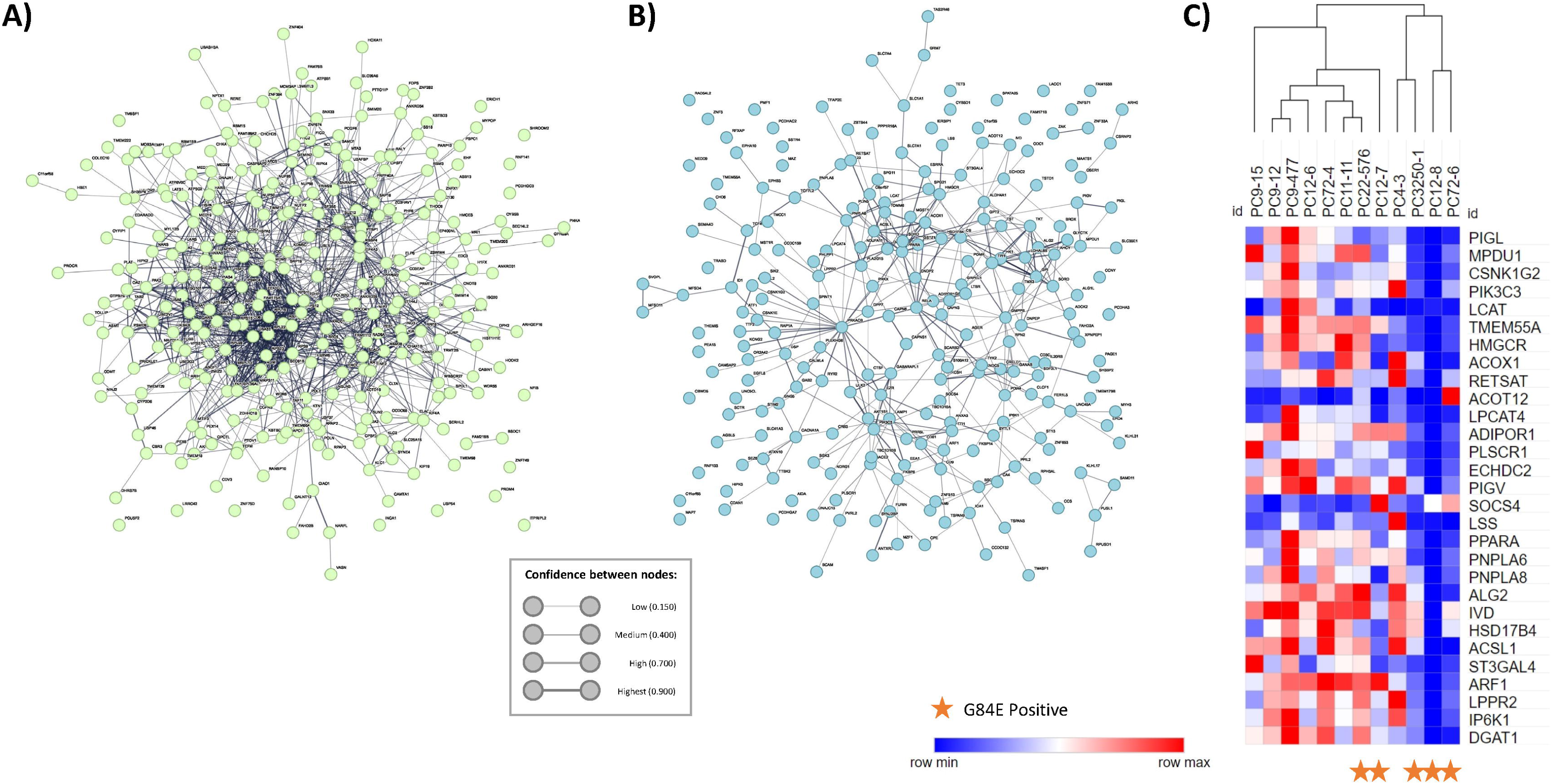
**A)** Protein-protein interaction web of DEG cluster one, which included genes involved in nucleic acid metabolic processes and transport, such as RNA and mRNA metabolic processes, translation and RNA splicing. **B)** Protein-protein interaction web of DEG cluster two which mapped to the cellular lipid metabolic process. **C)** Heatmap of the 30 lipogenesis pathway genes that mapped to the second DEG cluster as shown in Figure 2B. Heatmap represents raw gene expression data. *HOXB13* G84E positive tumours are depicted with a star.

### 3.2 Proteomic analysis of *HOXB13* G84E variant positive and wildtype prostate tumours

To explore whether there were detectable differences in protein expression between prostate tumours from G84E variant carriers and non-carriers, DIA-MS proteomic analysis was performed (28). In total, 2 429 proteins were quantified in 23 prostate tumour samples, including six *HOXB13* G84E variant positive and 17 wildtype tumours (Table 1; PC9-15 failed QC; PC12-03 and PC9-477 were excluded due to no viable cancer). Uniform manifold approximation and projection analyses revealed that samples clustered based on percentage of viable cancer in the tissue section but did not cluster on Gleason score (≤7 (3+4) *versus* ≥7 (4+3)) and age at diagnosis (<65 years *versus* ≥65 years) (Supplementary Figure 6).

We first examined whether HOXB13 protein expression was consistent with *HOXB13* gene expression. HOXB13 protein expression was on average slightly higher in G84E positive tumours (n=5) *versus* wildtype tumours (n=16), however this was not statistically significant (p=0.23; Figure 3A). This is consistent with our previous findings, where we observed no difference in expression in malignant cells from G84E variant carriers (n=9) and non-carriers (n=9) by immunohistochemistry (3).

**Figure 3.**
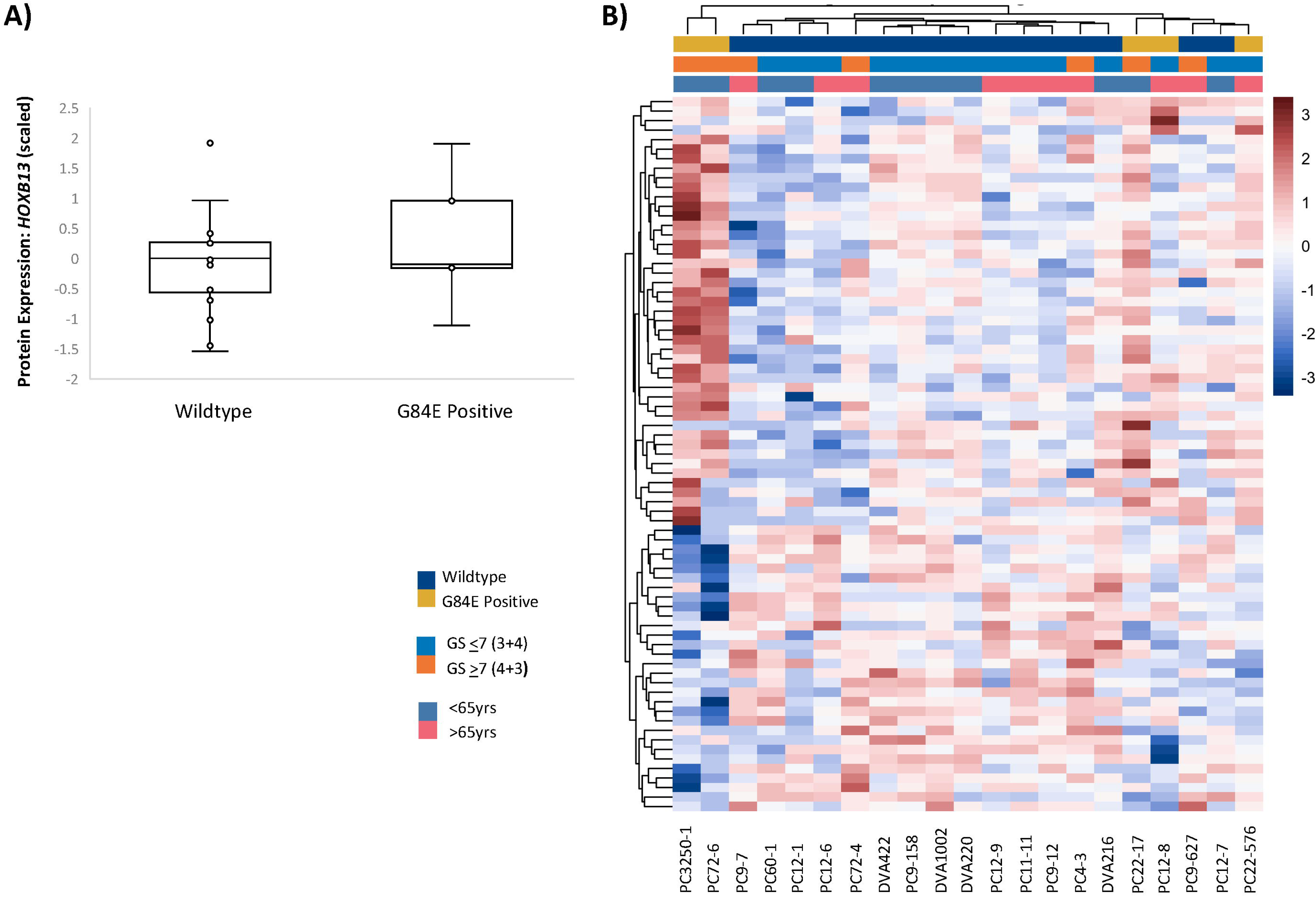
**A)** HOXB13 protein expression (scaled) of G84E positive (n=5) and wildtype tumours (n=16, p=0.23). Individual samples are labelled. **B)** Heatmap of the 132 proteins found to be differentially expressed between G84E positive and wildtype tumours when percentage of viable cancer was included as a covariate.

G84E variant positive and wildtype tumours revealed a 232-protein profile associated with G84E carrier status. A total of 103 proteins were found to be downregulated and 129 were upregulated in the tumours of *HOXB13* G84E variant carriers compared to non-carriers. Analysis was then repeated using percentage of cancer as a covariate (Table 1). This revealed a 132-protein profile associated with G84E carrier status, including 68 downregulated and 64 upregulated proteins (Figure 3B). Notably, 75 proteins overlapped this and the prior differential protein expression analysis. A full list of the DEPs can be found in Supplementary Tables 5 and 6.

GO analysis of the 75 overlapping proteins (30 downregulated and 45 upregulated) identified seven main biological processes (Supplementary Table 7). These included mitochondrial translation elongation (GO:0070125), purine nucleobase metabolic process (GO:0006144), fatty acid catabolic process (GO:0009062), purine nucleoside biphosphate metabolic process (GO:0034032), ribonucleoside bisphosphate metabolic process (GO:0033875), amide metabolic process (GO:0043603) and purine nucleotide metabolic process (GO:0006163). Cluster analysis revealed that the top cluster consisted of 23 proteins which mapped to lipogenesis pathways, including cellular lipid catabolic processes, fatty acid beta-oxidation and catabolic processes and short-chain fatty acid metabolic processes (Figure 4A; Supplementary Table 8). Notably, these processes were upregulated in tumours from G84E variant carriers (Figure 4C). A second cluster mapped to the desmosome, an intercellular junction that provides strong adhesion between cells (Figure 4B; Supplementary Table 8).

**Figure 4.**
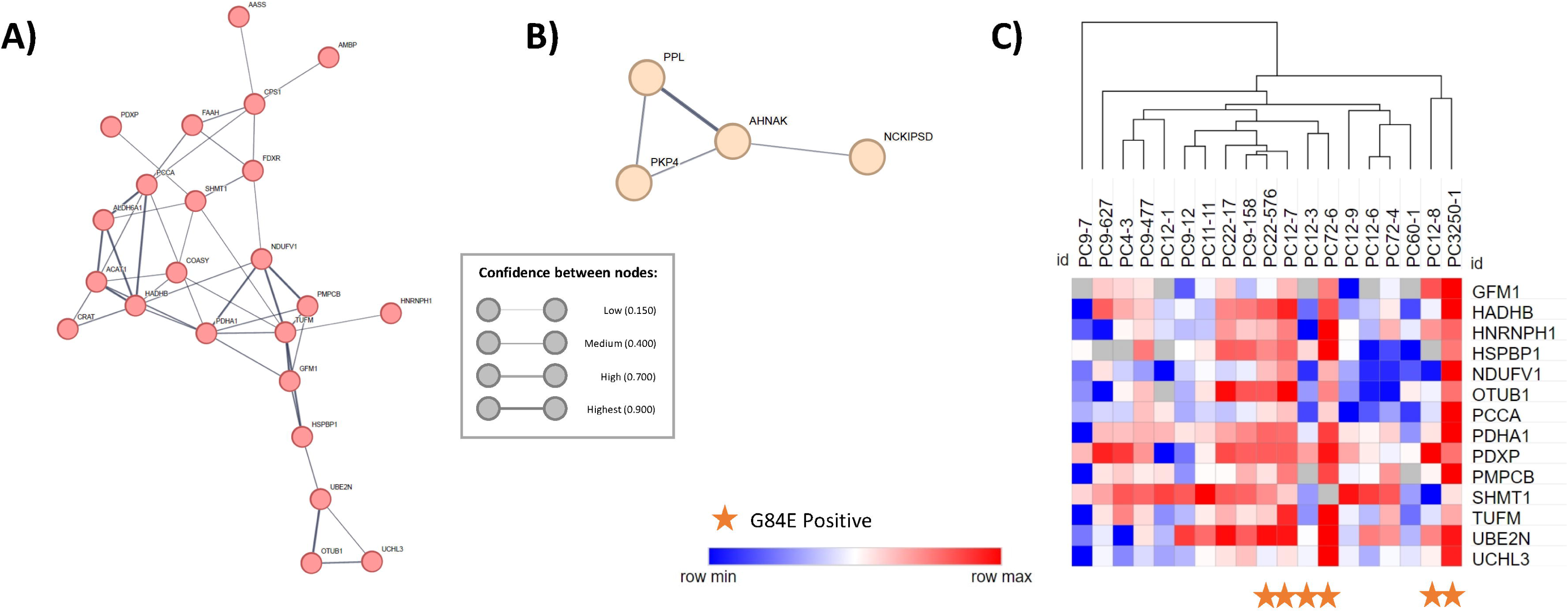
**A)** Protein-protein interaction web of DEP cluster one, which included proteins involved in mitochondrial translation, and fatty acid and lipid processes. **B**) Protein-protein interaction web of DEP cluster two which mapped to the desmosome, an intercellular junction that provides strong adhesion between cells. **C)** Heatmap of the 23 proteins in the top DEP cluster (as shown in Figure 4A). Heatmap represents raw protein expression data. *HOXB13* G84E positive tumours are depicted with a star.

### 3.3 A comparison of transcriptomic and proteomic data identifies three genes that are differentially expressed according to *HOXB13* G84E carrier status

A comparison of the 799 DEGs and 75 DEPs revealed three overlapping genes: *NCLN, NFIB* and *VASN*. All were downregulated in G84E variant positive tumours at the gene and protein level, except for *NCLN*, which was downregulated at the gene expression level and upregulated at the protein level (Figure 5A-B).

**Figure 5.**
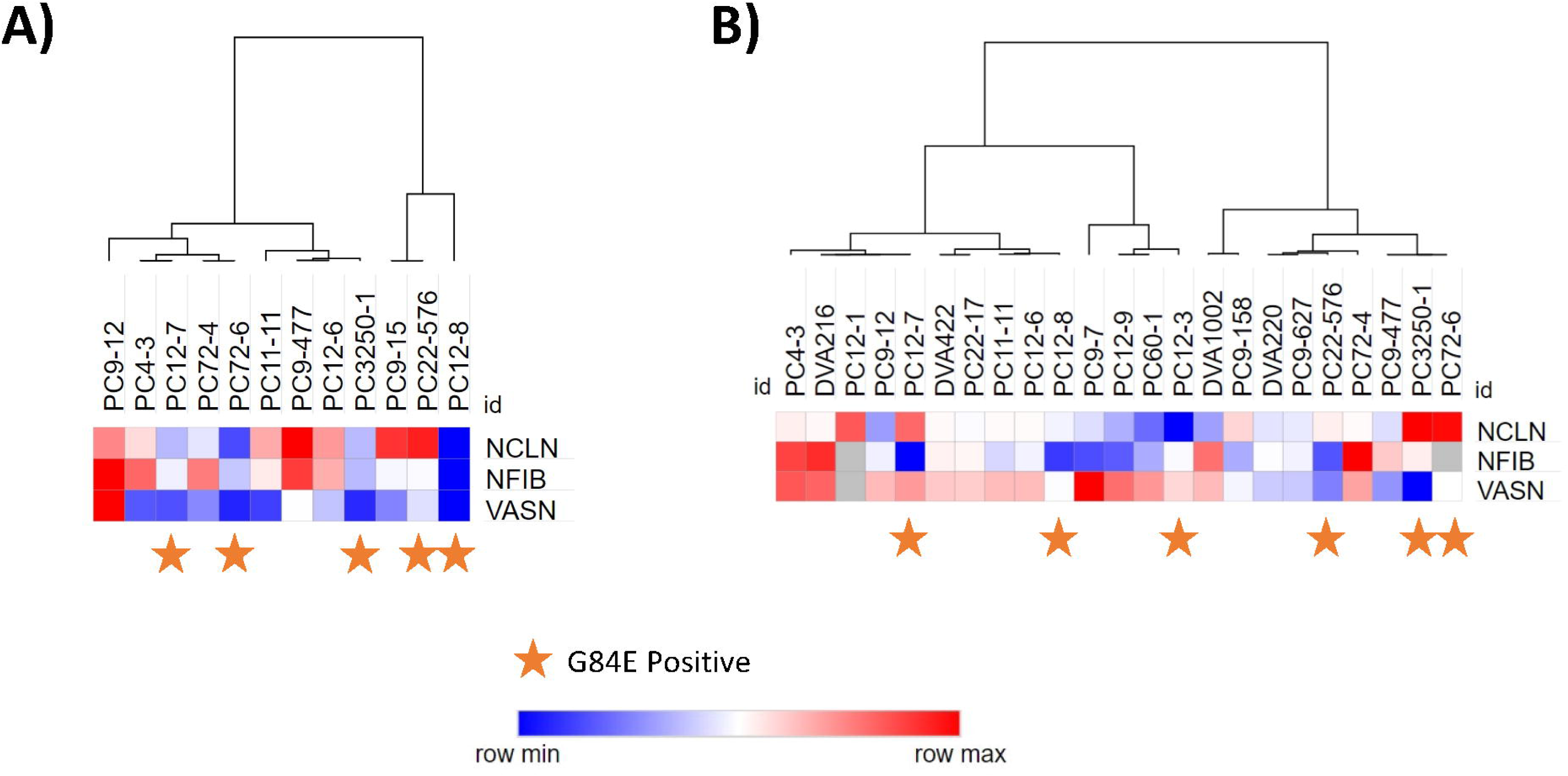
The G84E-like expression signature consists of the *NCLN*, *NFIB* and *VASN* genes. Raw gene expression data are shown in **A** and protein expression data in **B**.

Gene expression data from 493 TCGA tumours were then examined to identify tumours with a similar transcriptomic expression pattern (i.e., concurrent downregulation of *NCLN, NFIB* and *VASN*). In total, 52 of the 493 TCGA tumours were found to have a G84E-like expression signature (Figure 6A-C). While not statistically significant, the 52 G84E-like tumours were diagnosed at a slightly younger age (mean of 60 *vs.* 61 years) compared to the remainder of the dataset (n=441; p=0.17). Similarly, the G84E-like tumours tended to have lower grade disease (67% had GS ≤7) compared to the remainder of the dataset (58% had GS ≤7). Presence of the G84E variant has been hypothesised to cause a gain of function (2), and whilst the G84E variant is extremely rare, hypomethylation of the *HOXB13* gene, which is associated with higher gene expression, is commonly observed in primary prostate tumours (38). Examination of TCGA data revealed no difference in *HOXB13* methylation (mean of 0.048 *vs.* 0.045; p=0.11) or *HOXB13* gene expression (mean of 13,977 *vs* 13,509; p=0.23) between G84E-like samples and the remaining 441 tumours (Figure 6D-E).

**Figure 6.**
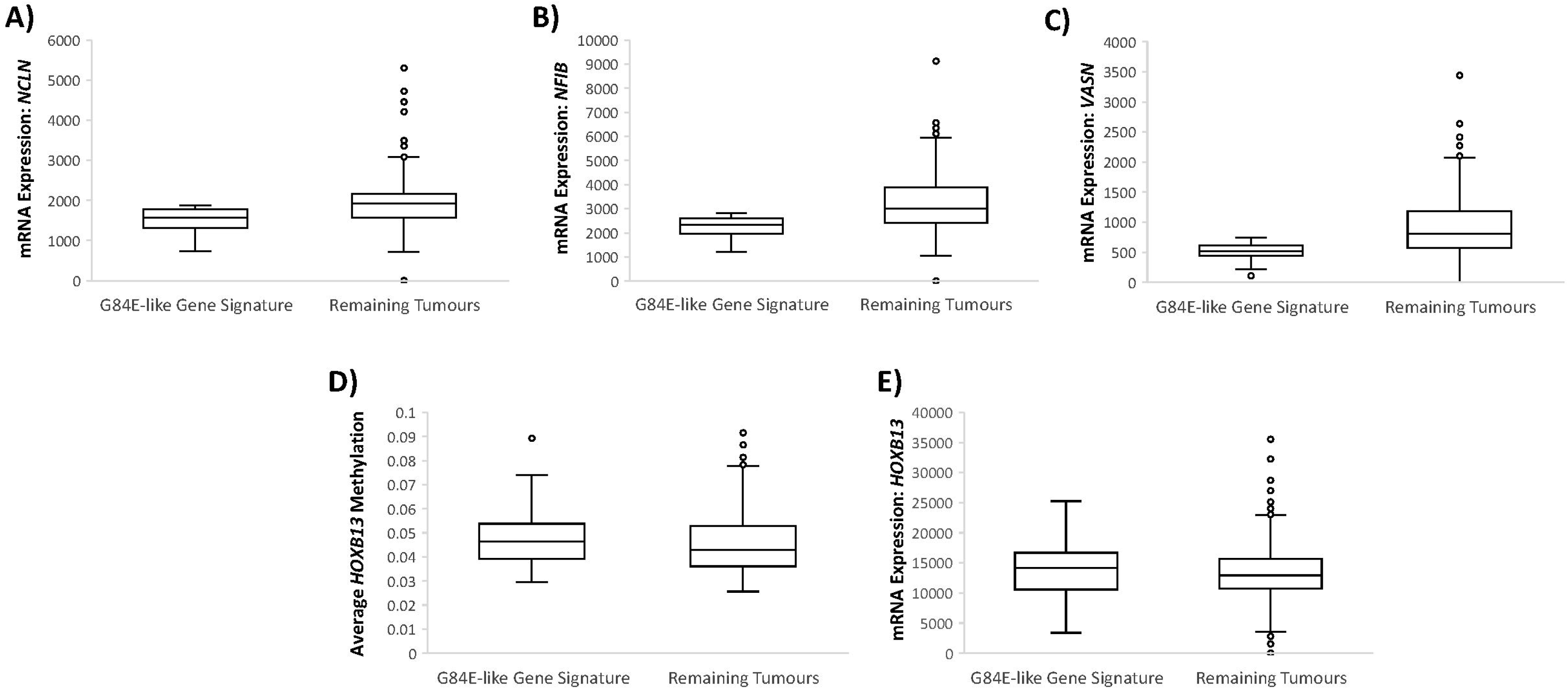
**A-C)** Gene expression of the *NCLN, NFIB* and *VASN* genes in TCGA tumours. In total, 52 tumours were found to have a similar expression pattern to our G84E positive tumours (i.e., concurrent downregulation of *NCLN, NFIB* and *VASN*, which are annotated as tumours with a G84E-like gene signature). **D)** *HOXB13* mRNA expression in tumours with the G84E-like gene signature (n=52) and the reminder of the TCGA dataset (n=441, p=0.23). **E)** Average *HOXB13* methylation in tumours with the G84E-like gene signature (n=52) and the reminder of the TCGA dataset (n=441, p=0.11).

## 4. DISCUSSSION

The molecular changes that occur in *HOXB13* G84E positive tumours have not previously been examined. To date, functionality of the G84E variant has only been examined using *in vitro* approaches. A study by Cardoso and colleagues (40) found no significant differences in tumourigenic characteristics of stably transfected cells overexpressing the G84E protein (compared to cells overexpressing wildtype HOXB13) (40), leaving unanswered questions regarding the role of G84E in tumour development. Computational modelling by Chandrasekaran and colleagues (2017) predicted that G84E increases HOXB13 protein stability, leading to an increased half-life with altered binding patterns and constitutive expression of downstream genes (39). Here, we used a proteogenomic approach to define the molecular changes occurring in *HOXB13* G84E variant positive tumours. The G84E variant was not associated with detectable differences in *HOXB13* expression at the gene and protein level in prostate tumours, consistent with our previous findings (3). Previously, we have shown that the variant allele is rarely detectable at the mRNA level in G84E positive prostate tumours and when present, is transcribed at lower levels than the wildtype allele (3).

In a novel approach, we sought to map the G84E tumour transcriptome and proteome and compare it to wildtype. A 799-gene and 75-protein profile associated with G84E carrier status was identified. Pathway analysis of these data highlighted 202 pathways in the transcriptome data and 57 pathways in the proteome data - the overlapping pathways comprising metabolic and cellular processes, amide biosynthesis and metabolic process, and multiple lipogenesis pathways. This novel finding in patient samples is consistent with recent work by Lu and colleagues (2022) (17) who showed that prostate cells expressing wildtype HOXB13 were able to repress lipogenic pathways through interaction with HDAC3 via modulation of DNA acetylation, whereas G84E variant expressing cells were unable to repress these pathways (17). Here, we provide the first evidence from patient samples that G84E positive tumours exhibit a similar proteome profile with upregulation of genes in the steroid metabolic, lipid biosynthesis and fatty acid metabolic pathways.

Integration of transcriptome and proteome data at the individual gene/protein level revealed three overlapping genes, including downregulation of *NCLN, NFIB* and *VASN* forming a G84E-like expression signature in variant positive tumours. Assessment of prostate tumours from TCGA revealed 52 tumours with a similar G84E-like expression signature. These 52 tumours had similar *HOXB13* methylation and expression compared to the remaining TCGA tumours. Whilst little is known about the role of these three genes in PrCa, two have established links to lipid metabolism pathways. *NFIB* is involved in the regulation of DNA binding activity and transcription by RNA polymerase II (41). Studies in hepatocytes have revealed that NFIB knockout is associated with perturbed lipid metabolism pathways (42), whereas increased expression of *NFIB* in PrCa has been found to correlate with *AR* expression *in vitro* (43). *VASN* is also implicated in hepatocyte autophagy and regulation of the normally tightly controlled glycogen/lipid metabolism. In relation to tumour development, VASN is involved in the negative regulation of epithelial to mesenchymal transition (EMT). One of the few studies in PrCa found that upregulation of *VASN* promotes EMT through a key regulator, the YAP/TAZ axis (44). VASN levels were measured in prostate tumours and serum, with their data suggesting that high VASN may have diagnostic value. Here, we found lower VASN expression in G84E positive tumours, which tend to exhibit low-grade disease, thus supporting this study by Cui and colleagues (44). Finally, *NCLN* enables ribosome binding activity, is involved in protein stabilisation and plays an important role in embryo development (37). Overall, additional *in vivo* and *in vitro* studies are required to establish and validate a causal link between the G84E variant and our observed expression signature.

The findings of our proteogenomic analysis are a first in PrCa, however there are several limitations to this study. Given the rarity of the G84E variant in the population (frequency of 0.8% in Europeans; gnomAD) and the absence of routine screening of individuals for this mutation, only six carriers had tumours available for this study, though this collection is one of very few. Insights into rare variant function have been generated previously using a similar sample size (25), and in other PrCa proteome studies (reviewed in Sadeesh *et al.,* 2021 (19)), recognising the inherent challenge in examining rare variants in human samples. Nevertheless, it is important that follow-up studies are performed in other cohorts of G84E positive tumours to confirm our findings. The absence of concordance between transcriptome and proteome data for the *NCLN* gene is noted, however, this discordance is a recognised phenomenon (45–49). Prior studies conducted in prostate tumours have shown that RNA expression is often a poor predictor of protein expression (50–52), with one study suggesting that mRNA abundance only explains around 10% of protein abundance (52). These studies highlight the importance of assessing the proteome in addition to inferring gene expression from the transcriptome. Furthermore, for this study, fresh tumour tissue was not available. Although the proteogenomic methodologies were optimised for FFPE and each tissue sample was obtained from the same pathology block, sections selected for RNA and protein analysis were not necessarily adjacent, thus will not be identical in composition due to tumour heterogeneity. PrCa is a heterogeneous, multifocal disease, which can include heterogeneity within a tumour (reviewed in Haffner, 2021) (53). Moreover, proteomic datasets do not represent the full proteome due to technical constraints. This results in fewer DEPs than DEGs, even when considering only protein coding genes, which leaves many proteins excluded from analysis. Notably, the utilisation of FFPE in this context allows us to perform research on samples available in the routine pathology setting. While FFPE presents technical challenges due to crosslinks formed between proteins and formaldehyde during sample processing (54), we have shown that our methodology adequately manages this potential problem. Furthermore, while RNA extracted from FFPE is often degraded, we have an established methodology to successfully generate high-quality transcriptome data from FFPE samples (26).

In conclusion, we report the first transcriptomes and proteomes generated from *HOXB13* G84E positive prostate tumours and, to the best of our knowledge, are the first to generate proteome data from familial PrCa tumours. Our 799-gene expression and 75-protein expression profiles highlighted critical shared pathways, including metabolic and cellular processes, amide biosynthesis and metabolic process, and significantly, numerous lipogenesis pathways. We also provide evidence from patient samples that G84E positive tumours exhibit a similar lipogenic proteome profile to that recently identified in an *in vitro* study by Lu and colleagues (2022) (17). Future studies are needed to confirm our findings in a larger collection of G84E variant positive tumours and clarify the role G84E plays in the perturbation of lipid metabolism pathways, utilising both *in vivo* and *in vitro* models. Further investigation of the molecular mechanisms involved will provide important insights into the diagnosis of PrCa and the development of novel therapeutic interventions for patients with the rare G84E high-risk variant.

## ACKNOWLEDGEMENTS

We are greatly indebted to the participants of our prostate cancer studies, the Tasmanian Cancer Registry staff, Tasmanian urologists, oncologists, pathologists and the wider Tasmanian clinical and research community for their ongoing support.

## AUTHORS’ CONTRIBUTIONS

KR and JRM contributed to sample preparation for both analyses. SW and PGH contributed to sample preparation and experiment for proteome analysis. KR and CJ contributed to the transcriptome data analysis, and ZN, ATA, PJR and QZ contributed to the proteome data analysis. SD and RLB reviewed the tumour samples. KR wrote the original draft and ZN, ATA, PGH, QZ, LMF and JLD contributed to the manuscript. LMF and JLD conceived the study, and KR, RRR and JLD led the study. All co-authors approved the final manuscript.

## ETHICS APPROVAL

This study was ethically approved by the University of Tasmania’s Human Research Ethics Committee (H0017040) and written informed consent was obtained from all participating individuals. For deceased cases, a waiver of consent was obtained to collected prostate tumour samples from pathology laboratories.

## DATA AVAILABILITY

Additional information can be found in the supplementary material. The mass spectrometry proteomics data have been deposited to the ProteomeXchange Consortium via the PRIDE (55) partner repository with the dataset identifier PXD051471.

