## Supplementary Material for "A proteogenomic approach to identifying a gene signature associated with *HOXB13* G84E carrier status in prostate cancer tumours"

**SUPPLEMENTARY INFORMATION**

**Variable window information for SWATH mass spectrometry:**

| | | |
|---|---|---|
| 399.5 | 406.5 | 5 |
| 405.5 | 412.5 | 5 |
| 411.5 | 418.5 | 5 |
| 417.5 | 424.5 | 5 |
| 423.5 | 430.5 | 5 |
| 429.5 | 436.5 | 5 |
| 435.5 | 442.5 | 5 |
| 441.5 | 448.5 | 5 |
| 447.5 | 454.5 | 5 |
| 453.5 | 459.5 | 5 |
| 458.5 | 464.5 | 5 |
| 463.5 | 469.5 | 5 |
| 468.5 | 474.5 | 5 |
| 473.5 | 479.5 | 5 |
| 478.5 | 484.5 | 5 |
| 483.5 | 489.5 | 5 |
| 488.5 | 494.5 | 5 |
| 493.5 | 499.5 | 5 |
| 498.5 | 504.5 | 5 |
| 503.5 | 509.5 | 5 |
| 508.5 | 514.5 | 5 |
| 513.5 | 519.5 | 5 |
| 518.5 | 524.5 | 5 |
| 523.5 | 529.5 | 5 |
| 528.5 | 534.5 | 5 |
| 533.5 | 539.5 | 5 |
| 538.5 | 544.5 | 5 |
| 543.5 | 549.5 | 5 |
| 548.5 | 554.5 | 5 |

| | | |
|---|---|---|
| 553.5 | 559.5 | 5 |
| 558.5 | 564.5 | 5 |
| 563.5 | 569.5 | 5 |
| 568.5 | 574.5 | 5 |
| 573.5 | 579.5 | 5 |
| 578.5 | 584.5 | 5 |
| 583.5 | 589.5 | 5 |
| 588.5 | 594.5 | 5 |
| 593.5 | 599.5 | 5 |
| 598.5 | 604.5 | 5 |
| 603.5 | 609.5 | 5 |
| 608.5 | 614.5 | 5 |
| 613.5 | 619.5 | 5 |
| 618.5 | 624.5 | 5 |
| 623.5 | 629.5 | 5 |
| 628.5 | 634.5 | 5 |
| 633.5 | 639.5 | 5 |
| 638.5 | 644.5 | 5 |
| 643.5 | 649.5 | 5 |
| 648.5 | 654.5 | 5 |
| 653.5 | 660.5 | 5 |
| 659.5 | 666.5 | 5 |
| 665.5 | 672.5 | 5 |
| 671.5 | 678.5 | 5 |
| 677.5 | 684.5 | 5 |
| 683.5 | 690.5 | 5 |
| 689.5 | 696.5 | 5 |
| 695.5 | 702.5 | 5 |
| 701.5 | 708.5 | 5 |
| 707.5 | 714.5 | 5 |
| 713.5 | 720.5 | 5 |

| | | |
|---|---|---|
| 719.5 | 726.5 | 5 |
| 725.5 | 732.5 | 5 |
| 731.5 | 738.5 | 5 |
| 737.5 | 744.5 | 5 |
| 743.5 | 750.5 | 5 |
| 749.5 | 756.5 | 5 |
| 755.5 | 763.5 | 5 |
| 762.5 | 770.5 | 5 |
| 769.5 | 777.5 | 5 |
| 776.5 | 784.5 | 5 |
| 783.5 | 791.5 | 5 |
| 790.5 | 798.5 | 5 |
| 797.5 | 805.5 | 5 |
| 804.5 | 812.5 | 8 |
| 811.5 | 819.5 | 8 |
| 818.5 | 826.5 | 8 |
| 825.5 | 834.5 | 8 |
| 833.5 | 842.5 | 8 |
| 841.5 | 850.5 | 8 |
| 849.5 | 858.5 | 8 |
| 857.5 | 867.5 | 8 |
| 866.5 | 876.5 | 8 |
| 875.5 | 885.5 | 8 |
| 884.5 | 894.5 | 8 |
| 893.5 | 903.5 | 8 |
| 902.5 | 914.5 | 8 |
| 913.5 | 925.5 | 8 |
| 924.5 | 936.5 | 8 |
| 935.5 | 950.5 | 8 |
| 949.5 | 964.5 | 8 |
| 963.5 | 978.5 | 8 |

| | | |
|---|---|---|
| 977.5 | 992.5 | 8 |
| 991.5 | 1011.5 | 10 |
| 1010.5 | 1030.5 | 10 |
| 1029.5 | 1054.5 | 10 |
| 1053.5 | 1078.5 | 10 |
| 1077.5 | 1117.5 | 10 |
| 1116.5 | 1156.5 | 10 |
| 1155.5 | 1200.5 | 10 |
| 1199.5 | 1249.5 | 10 |

**SUPPLEMENTARY FIGURES**

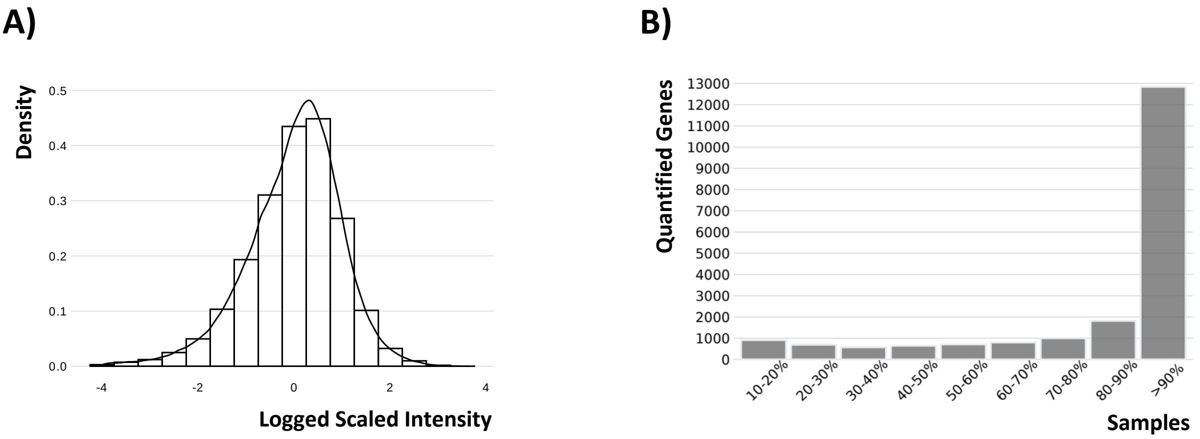


**Supplementary Figure 1.** Intensity distribution of the AmpliSeq data (A) and genes quantified in fraction of samples (B). Supplementary Figure 1B shows that ~13,000 genes (from the total number of quantified genes) were quantified in >90% of samples. Thus, a large proportion of total quantifiable genes were available for analysis in a large set of samples.

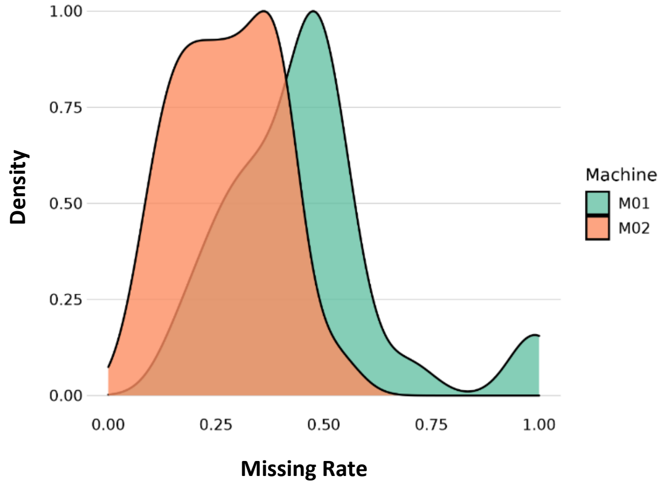


**Supplementary Figure 2.** Missing rate per individual on each machine (M1, M2) in the non-normalized peptide matrix.

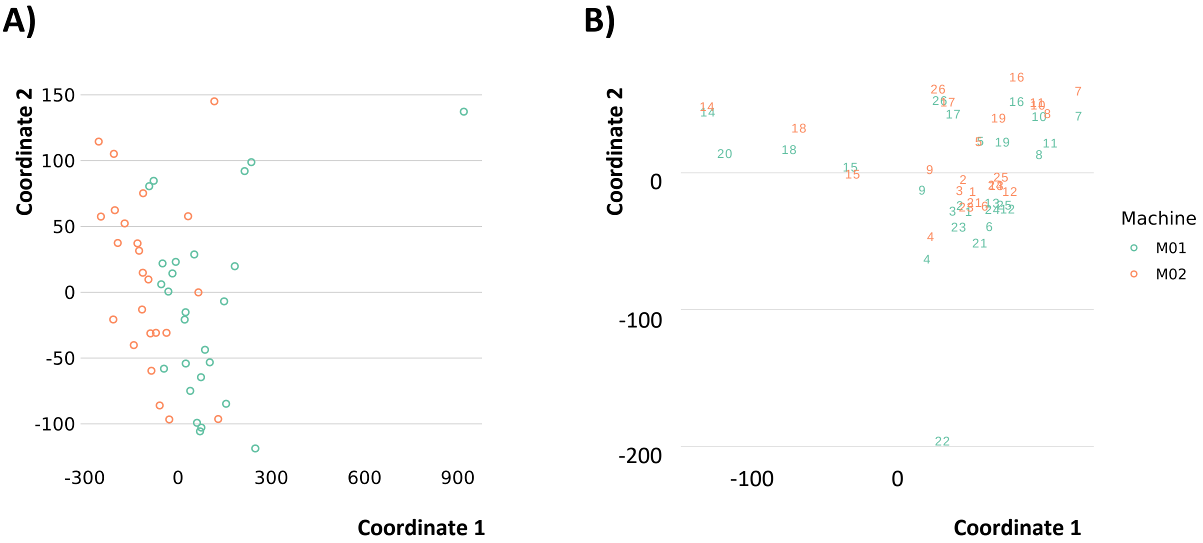


**Supplementary Figure 3.** MDS plot of non-normalised (A) and normalised peptides (B).

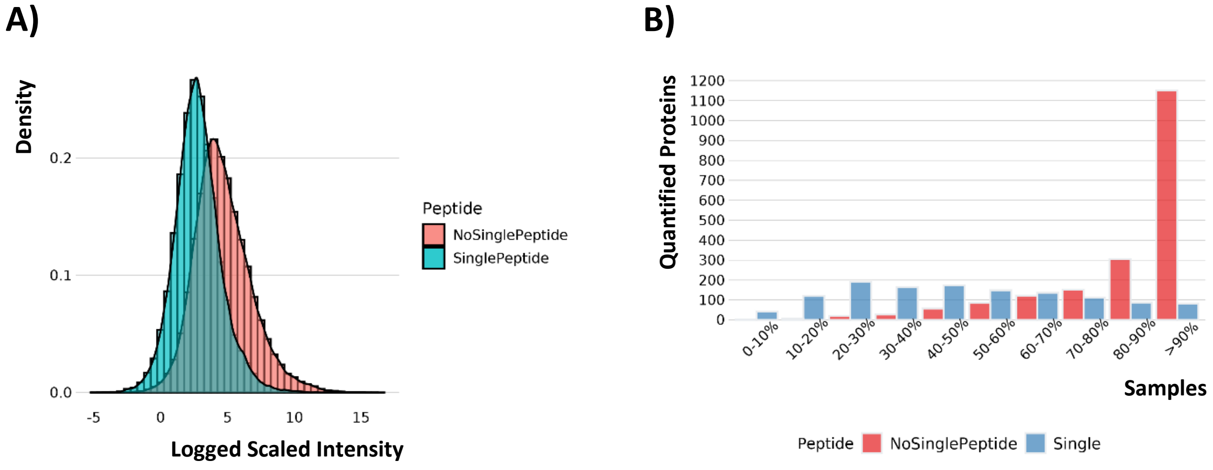


**Supplementary Figure 4.** Standardised matrix intensity distribution of the proteome data (A) and proteins quantified in fraction of samples (B).

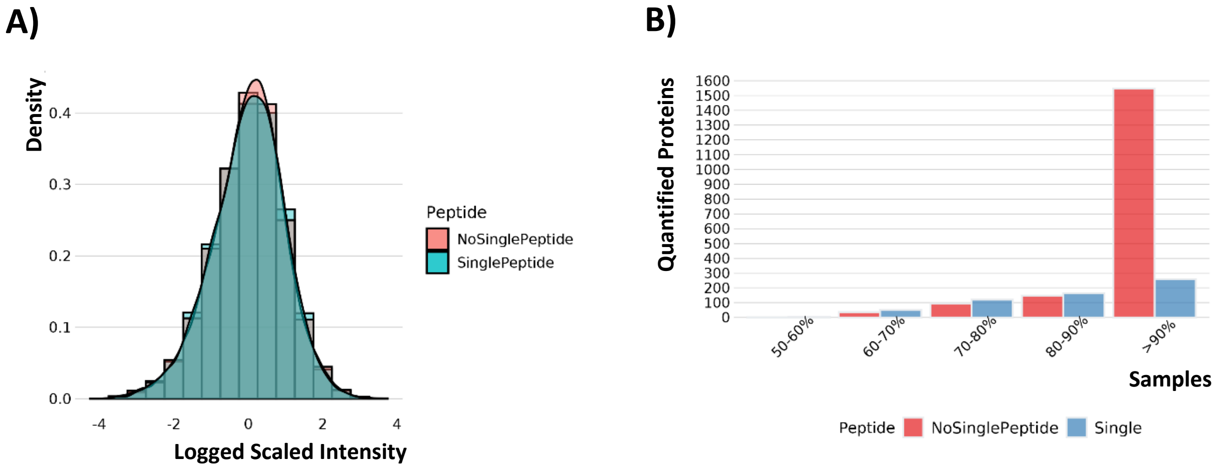


**Supplementary Figure 5.** Standardised matrix intensity distribution of the DIA-NN protein data (A) and proteins quantified in fraction of samples (B).

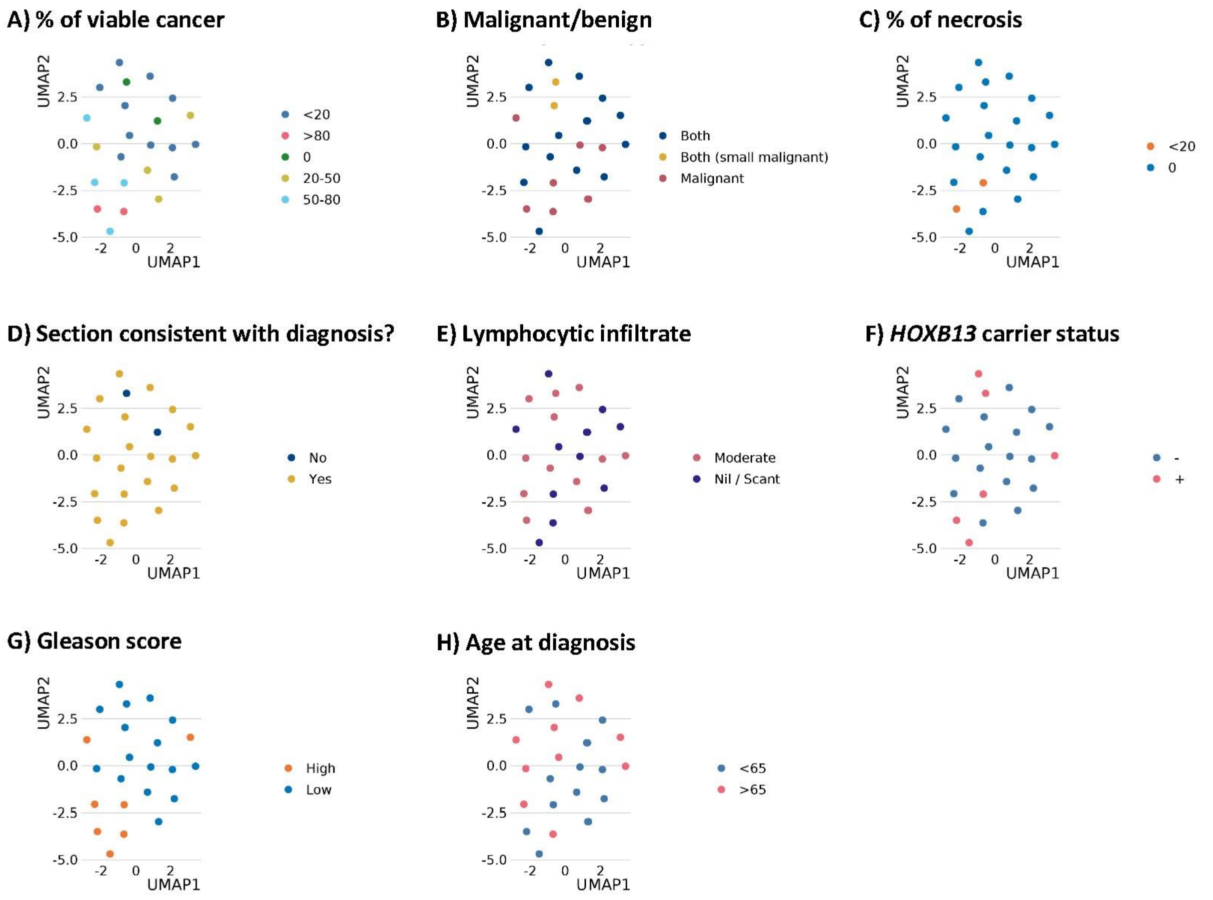


**Supplementary Figure 6.** UMAP of the filtered protein matrix showing clustering of proteomes based on percentage (%) of viable cancer (A), malignant/benign regions in tumour section (B), % of necrosis (C), assayed section consistent with diagnosis (D), lymphocytic infiltrate (E), *HOXB13* G84E carrier status; -: wildtype, +: G84E positive (F), Gleason score; high: GS ≥7 (4+3), low: GS ≤7 (3+4) (G), age at diagnosis (H).
